# Social Frailty Index: Development and Validation of an Index of Social Attributes Predictive of Mortality in Older Adults

**DOI:** 10.1101/2022.05.24.22275541

**Authors:** Sachin J Shah, Sandra Oreper, Sun Young Jeon, W. John Boscardin, Margaret Fang, Kenneth E Covinsky

## Abstract

**Objective:** To develop and validate the Social Frailty Index, a summary measure of social risk in older adults, and determine its ability to risk stratify beyond traditional medical risk models

**Design:** Prognostic model development and validation using demographics and a comprehensive inventory of social characteristics

**Setting:** The Health and Retirement Study, a longitudinal, nationally representative survey of U.S. adults >50 years. We developed the model using the 2010 wave and validated it using the 2012 wave; there was no overlap between 2010 and 2012 respondents.

**Participants:** 8250 adults aged ≥65 years who completed the Psychosocial and Lifestyle Questionnaire (4302 in the 2010 development cohort, 3948 in the 2012 validation cohort).

**Main exposure:** Demographic and social characteristics

**Main outcome:** 4-year mortality

**Results:** Within 4 years of the baseline interview, 22% of study participants in both the development and validation cohort had died. Drawn from 183 possible predictors, the final model included age, gender, and 8 social predictors: lacking neighborhood cleanliness, low perceived control over financial situation, having children and meeting with them less than yearly, not working for pay, less active with children (grandchildren, neighborhood, nieces/nephews), no volunteering or charity work, feeling isolated from others, being treated with less courtesy or respect. In the validation cohort, the model discriminated well (c-statistic 0.73) and was strongly associated with 4-year mortality (7.3% in the lowest decile, 59.9% in the highest decile). Also, the Social Frailty Index meaningfully risk-stratified participants beyond the Charlson score, a commonly used medical comorbidity index, and the Lee Index score, a comorbidity and function model.

**Conclusion:** This prognostic index, which includes age, gender, and 8 social characteristics, accurately risk stratifies older adults and refines the prediction of commonly used comorbidity- and function-based risk models.

## INTRODUCTION

Risk prediction and prognostication are core to clinical medicine, medical research, and healthcare policy. For example, life expectancy informs benefits and harms of cancer screening, baseline risk measurement is central to observational research, and risk adjustment is crucial in quality measures.^1–3^ Traditional approaches to risk prediction rely heavily on measuring the prognostic impact of medical comorbidities.^3–5^ These efforts have yielded widely-used summary measures of medical risk.

However, a rich literature also demonstrates that a wide range of social factors meaningfully predict health outcomes.^6–8^ For example, social support predicts reduced rates of nursing home stays,^9^ loneliness portends higher rates of functional decline and death,^10^ and social network strength is associated with lower rates of cognitive decline.^11^

Although social attributes are predictive of key aging outcomes, we lack an efficient way to summarize the prognostic impact of social factors. Practicality is a key impediment—for use in a clinical, research, or policy setting, a social risk model is more likely to be implemented if it is easy to use. The few existing social risk models that have been developed are expansive inventories.^12–15^ These models are comprehensive and predictive, albeit unwieldy to implement. Some have sought to address usability by relying exclusively on area-level data. Such efforts produced the Center for Disease Control’s Social Vulnerability Index,^16^ the Area Deprivation Index,^17^ and the English Indicies of Deprivation^18^, which are useful for area-level interventions and planning.

However, such measures cannot assess an individual’s risk because inferences from group-level analyses cannot be reliably applied to individuals within those groups, a principle known as ecological fallacy.^19^ Codifying which social elements to include is challenging; as a result, many studies exclude social risk factors altogether and, in doing so, run the risk of biased measurement.^20, 21^

The primary aim of this study was to create the Social Frailty Index, a parsimonious person-level social risk prediction model derived from a comprehensive inventory of social characteristics that predicts mortality in older adults. Our goal was to identify a small subset of risk factors that reflects social risk, not to identify causal factors or all possible social risk factors. Our second aim was to determine if the Social Frailty Index improves risk stratification beyond existing risk models.

## METHODS

### Study participants

We developed a longitudinal cohort of older adults from the Health and Retirement Study (HRS) to develop the Social Frailty Index. The HRS is a longitudinal, nationally representative survey of more than 43,000 Americans aged 50 years and older.^22, 23^ The goal of the HRS is to measure changes in health, wealth, social structure, and function as participants age. Participants are interviewed every 2 years by phone, in person or via internet surveys. We included participants who were 65 years and older who completed the Psychosocial and Lifestyle Questionnaire in 2010 or 2012. The 2010 cohort was used to develop the model, and the 2012 cohort was used to validate the model. Because the questionnaire is administered to a random half of the total HRS cohort every two years, there is no overlap between 2010 and 2012 respondents.

### Social predictors

We identified predictors using the Social Frailty in Older Adults framework (**Appendix 1**), a framework articulated by sociologists and gerontologists based on Social Production Function theory—it conceptualizes social frailty as “a lack of resources to fulfill one’s basic social needs.”^24^ The framework identifies 4 social domains relevant to aging—General Resources, Social Resources, Social Activities, Fulfillment of Basic Social Needs. Using the HRS core interview and the Psychosocial and Lifestyle Questionnaire, we identified 472 potential social predictors. Two investigators (SJS, SO) independently reviewed each predictor to determine if it fit within the Social Frailty in Older Adults framework. Any differences were reconciled through consensus discussion. This process yielded 183 candidate predictors (**Appendix 2**).

### Outcome

Our primary outcome was all-cause mortality 4 years after the interview, mirroring the time horizon used in prior mortality prediction studies.^4^ Mortality was determined using a combination of the National Death Index and HRS surviving family member exit interviews. We identified 3 secondary outcomes, all assessed over 4 years—loss of activity of daily living (ADL) independence, prolonged nursing home stay, and hospitalization. Loss of ADL independence was defined as being fully ADL independent at baseline interview and requiring help with one or more ADLs at 4 years. Nursing home use was defined as spending 90 nights or more in a nursing home within 4 years of the interview. Hospitalization was defined as self-reported hospital admission lasting 2 nights or more.

### Development and validation of the primary model

We used the 2010 cohort to derive the prediction model. We determined the functional form of continuous predictors by assessing linear, log transformation, and exponential transformation against the primary outcome (mortality at 4 years). We selected the functional form with the lowest Bayesian information criterion (BIC). We determined the functional form of ordinal predictors by assessing linear, categorial, and manual categorization against the primary outcome selecting the functional form with the lowest BIC. Missing predictors were imputed using single imputation. Categorical variables were imputed with the mode value and continuous variables were imputed with the median value.

We used a two-step procedure to derive the prediction model.^25^ First, from the set of 183 predictors, we used LASSO regression with a lambda parameter chosen by 10-fold cross-validation to produce the smallest subset of predictors. Then, we used the selected predictors to fit a logistic regression model that estimated the 4-year risk of death. In this step, we removed several additional to improve parsimony and face validity and reduce collinearity. Removing these predictors improved the model BIC and only marginally affected the AUC (0.74 to 0.73). This final model was then validated in the 2012 cohort. In the validation cohort, we determined discrimination and calibration. We also determine the model’s discriminatory capacity for secondary outcomes. In the 2012 cohort, we determined if the Social Frailty Index could further stratify participants’ mortality risk beyond the Lee Index and beyond the Charlson score in the subset with Medicare claims linkage.^3, 4^

We report all results with 95% confidence intervals or two-sided P value. The a priori significance threshold was P < .05. We performed analyses using SAS 9.4 (Cary, NC) and R 4.0.3 (Vienna, Austria). The TRIPOD checklist can be found in Appendix 3.^26^

## RESULTS

### Baseline characteristics

4302 participants were included in the 2010 development cohort (**Appendix 4**). The development cohort’s median age was 75 years, and 57% were women (**Table 1**). Regarding health and function, 27% reported fair or poor health, 56% reported ever using tobacco, and 26% had a screening test consistent with cognitive impairment or dementia. Within 4 years of the baseline interview, 22% of study participants in both the development cohort (960/4302) and validation cohort (882/3948) had died.

**Table 1:** Cohort characteristics, demographics, health status, and function. IQR – interquartile range; ADL – activities of daily living

|  | 2010<br>Development<br>cohort<br>(n=4302) | 2012<br>Validation<br>cohort<br>(n=3948) |
| --- | --- | --- |
| **Demographics** |  |  |
| Age, years, (median [IQR]) | 75 [70, 80] | 75 [71, 80] |
| Women (%) | 2444 (57) | 2308 (58) |
| **Health Status** |  |  |
| Fair or poor self-reported health (%) | 1169 (27) | 1116 (28) |
| Significant pain (%) | 240 (6) | 211 (5) |
| Diabetes (%) | 1033 (24) | 1020 (26) |
| Cancer (excluding minor skin cancer) (%) | 907 (21) | 850 (22) |
| Heart disease (%) | 1350 (31) | 1323 (34) |
| Stroke (%) | 465 (11) | 478 (12) |
| Arthritis (%) | 2973 (69) | 2797 (71) |
| Depression (%) | 489 (12) | 474 (12) |
| Ever tobacco use (%) | 2398 (56) | 2219 (57) |
| **Function** |  |  |
| Help with any ADL (%) | 377 (9) | 375 (10) |
| Cognition (%) |  |  |
| Dementia | 266 (6) | 270 (7) |
| Impairment, not dementia | 870 (20) | 813 (21) |
| Intact | 3166 (74) | 2865 (73) |
| Visual impairment (%) | 952 (22) | 917 (23) |
| Hearing impairment (%) | 1113 (26) | 1039 (26) |

In the domain of General Resources and Life History, 19% had less than a high school education, and 27% were military veterans (**Table 2**). Regarding Social Resources, 62% were married or partnered, 92% had living children, and 64% reported they “often have someone they can talk to.” In Social Activities, 6% reported meeting with their children less than yearly, and 65% reported writing or emailing friends monthly or more frequently. Concerning Fulfillment of Basic Social Needs, 5% reported often feeling isolated from others, 76% reported being rarely being treated with less courtesy or respect than other people (never or less than once a year), and 12% reported not having a major activity such as a job, looking after the home, or volunteer work.

**Table 2:** Cohort social characteristics

|  | 2010<br>Development<br>cohort<br>(n=4302) | 2012<br>Validation<br>cohort<br>(n=3948) |
| --- | --- | --- |
| **General Resources and Life History** |  |  |
| Race (%) |  |  |
| Black | 500 (12) | 485 (12) |
| White | 3665 (85) | 3315 (84) |
| Hispanic ethnicity (%) | 278 (6) | 321 (8) |
| Local area is full of rubbish and litter* | 733 (18) | 755 (20) |
| Low self-reported control over financial situation** (%) | 441 (10) | 402 (11) |
| Military veteran (%) | 1152 (27) | 968 (25) |
| Satisfaction with daily life and leisure activities (1 to 5 scale) (median [IQR]) ^ | 2 [1, 3] | 2 [1, 3] |
| Not at all satisfied with retirement (%) | 192 (5) | 181 (5) |
| Education (%) |  |  |
| Less than high school | 834 (19) | 829 (21) |
| High school | 1685 (39) | 1459 (37) |
| Some college | 897 (21) | 860 (22) |
| College and above | 885 (21) | 800 (20) |
| **Social Resources** |  |  |
| Married or Partnered (%) |  |  |
| Married or partnered | 2666 (62) | 2353 (60) |
| Never married | 107 (2) | 103 (3) |
| Separated or divorced | 374 (9) | 398 (10) |
| Widowed | 1155 (27) | 1094 (28) |
| Living children (%) | 3799 (92) | 3367 (92) |
| Has friends (%) | 3877 (93) | 3481 (91) |
| Number of living siblings (median [IQR]) | 2 [1, 3] | 2 [1, 4] |
| Often has someone they can talk to (%) | 2690 (64) | 2453 (63) |
| **Social Activities** |  |  |
| Presently working for pay (%) | 738 (18) | 630 (16) |
| Activities with grandchildren, niece/nephews or neighborhood children (median [IQR]) ^^ | 5 [4, 7] | 5 [4, 7] |
| Does no volunteering or charity work (%) | 2205 (53) | 2163 (56) |
| Goes to a sport, social, or other club (median [IQR]) \$ | 6 [5, 7] | 6 [5, 7] |
| Meets with children less than once a year (%) | 246 (6) | 276 (7) |
| Writes or emails friends monthly or more frequently (%) | 2581 (65) | 2348 (64) |
| **Fulfillment of Basic Social Needs** |  |  |
| Feel isolated from others (%) |  |  |
| Often | 211 (5) | 194 (5) |
| Some of the time | 1013 (24) | 1018 (26) |
| Hardly ever or never | 2948 (69) | 2600 (66) |
| Often feel in tune with the people around you (%) | 1990 (48) | 1814 (47) |
| Feel part of a group of friends |  |  |
| Often | 2023 (48) | 1817 (47) |
| Some of the time | 1593 (38) | 1430 (37) |
| Hardly ever or never | 572 (14) | 602 (16) |
| You are treated with less courtesy or respect than other people (never or less than once a year) (%) | 3209 (76) | 2923 (76) |
| People act as if they think you are not smart (never or less than once a year) (%) | 3336 (79) | 3068 (80) |

### Model development results

The model development procedure yielded a model of 10 predictors from all 4 domains in the Social Frailty in Older Adults Framework (**Table 3**). The 10 predictors include age and gender, 2 measures of General Resources and Life Events (neighborhood cleanliness; perceived control over their financial situation), 1 measure of Social Resource and Social Activities (has children and meets with them less than yearly), 3 measures of Social Activities (working for pay; less active with grandchildren, neighborhood children, nieces/nephews; no volunteering or charity work), and 2 measures reflecting Fulfillment of Basic Social Needs (feeling isolated from others; being treated with less courtesy or respect).

**Table 3:**
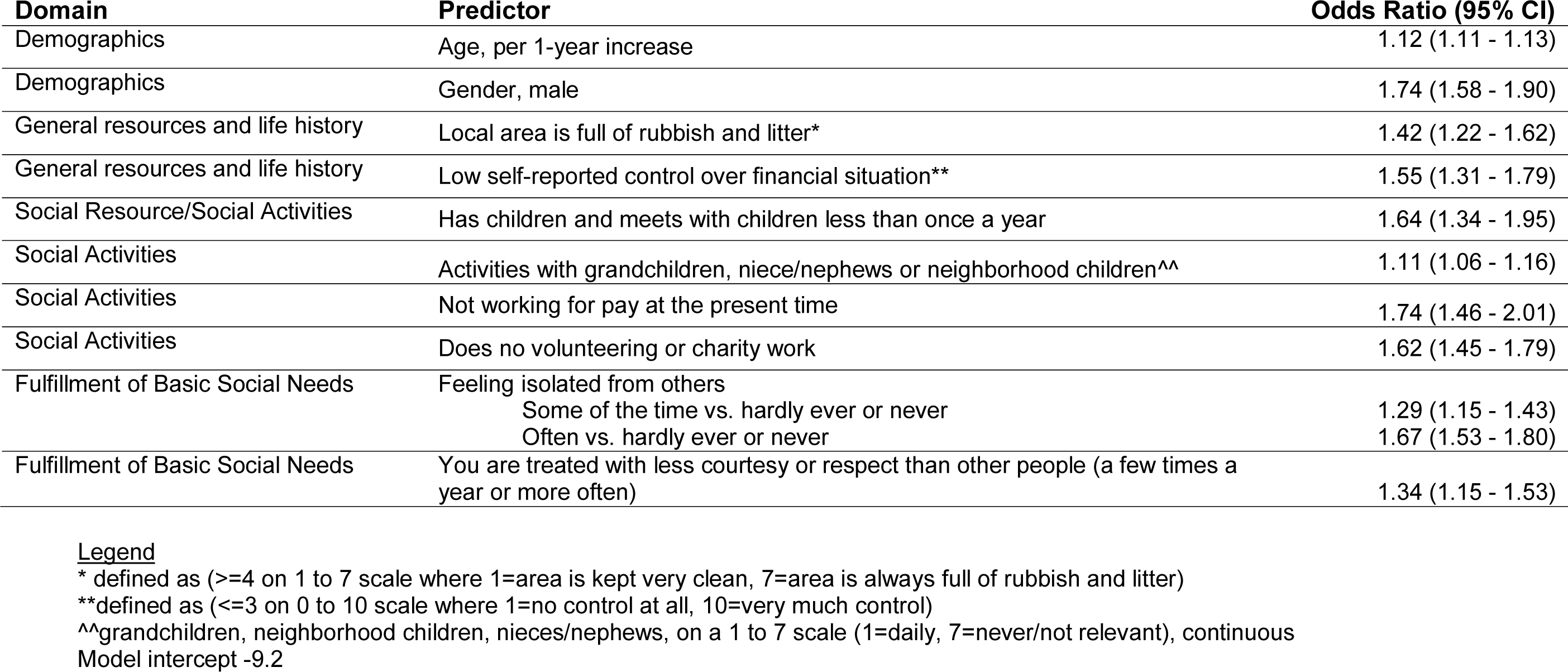
Individual Predictors in the Social Frailty Index

Beyond age and gender, which are commonly used in prediction models, in the final model, the most prominent predictors of death include not working for pay (OR 1.74, 95% CI 1.46 to 2.01), meeting with children less than once a year (OR 1.64, 95% CI 1.34 to 1.95), and often feeling isolated from others (OR 1.67, 95% CI 1.53 to 1.80).

### Validation measures

The Social Frailty Index performed well in the 2012 validation cohort. The model was well calibrated (**Appendix 5**), observed and expected mortality in the validation data were highly correlated. In the lowest decile, the observed 4-year mortality was 7.3% (predicted 4.4%), and in the highest decile was 59.9% (predicted 62.2%). The model discriminated well with an AUC of 0.76 to predict death in the development cohort and 0.73 in the validation cohort (**Appendix 6**). The model also performed well when used to predict new ADL dependence (AUC 0.72) and nursing home stays (AUC 0.74). The model performed modestly when used to predict hospitalization (AUC 0.64).

### Risk stratification beyond Charlson Score

The Social Frailty Index meaningfully risk-stratified participants beyond the Charlson score, a commonly used medical comorbidity index. The Social Frailty Index and the Charlson score are weakly correlated (Pearson’s correlation of 0.17, CI 0.12- 0.21, p<0.001) in a subset of the validation cohort who have 12 months of Medicare claims data before their baseline interview (2226 of 3948). **Figure 1** illustrates that the Social Frailty Index risk stratifies beyond the Charlson score in all 3 tertiles of the Charlson score. For example, in the validation cohort’s highest tertile of Charlson score, the observed mortality rate with a high Social Frailty Index score was 47% vs. 30% in the low Social Frailty tertile (p<0.001).

**Figure 1:**
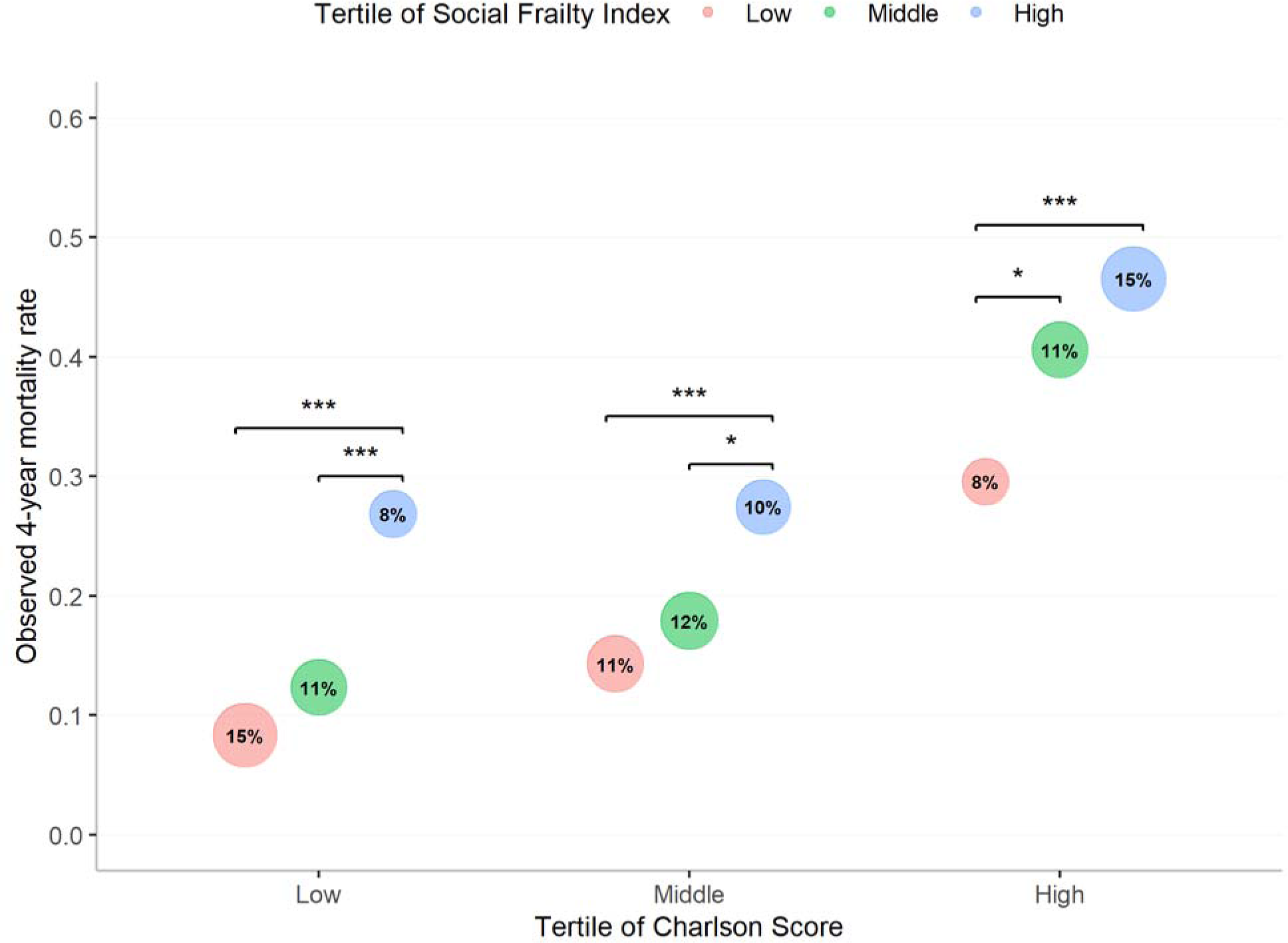
Observed mortality in 2012 Validation Cohort by Social Frailty and Charlson score The bubble chart compares observed mortality in the validation cohort by tertile of Social Frailty within tertiles of Charlson score, a comorbidity risk model. The Charlson score cohort was completed in a subset of the study cohort where 12 months of Medicare data were available to calculate a Charlson score (2226 of 3948). Since the Charlson score does not include age, when comparing it with the Social Frailty Index, we remove age from the Social Frailty Index to provide a fair comparison. The area of each bubble is proportional to the total validation cohort that falls the specific group (e.g., 15% of the cohort has a low Charlson score and low Social Frailty Index score). Significantly different values are highlighted by a bracket. *** p<0.001, ** p<0.01, * p<0.05. Results presented in tabular form in Appendix 7.

### Risk stratification beyond Lee Index

The Social Frailty Index stratified participants’ risk beyond the Lee Index, a commonly used model that uses comorbidities and function to predict mortality. The Social Frailty Index and the Lee Index were modestly correlated (Pearson’s correlation of 0.63, CI 0.61-0.65, p<0.001). **Figure 2** illustrates that the Social Frailty Index risk stratifies beyond the Lee Index, specifically those with Middle or High Lee Index scores. For example, in the highest tertile of Lee score in the validation cohort, the observed mortality rate with a high Social Frailty Index score was 51% vs. 29% in the low Social Frailty tertile (p<0.001).

**Figure 2:**
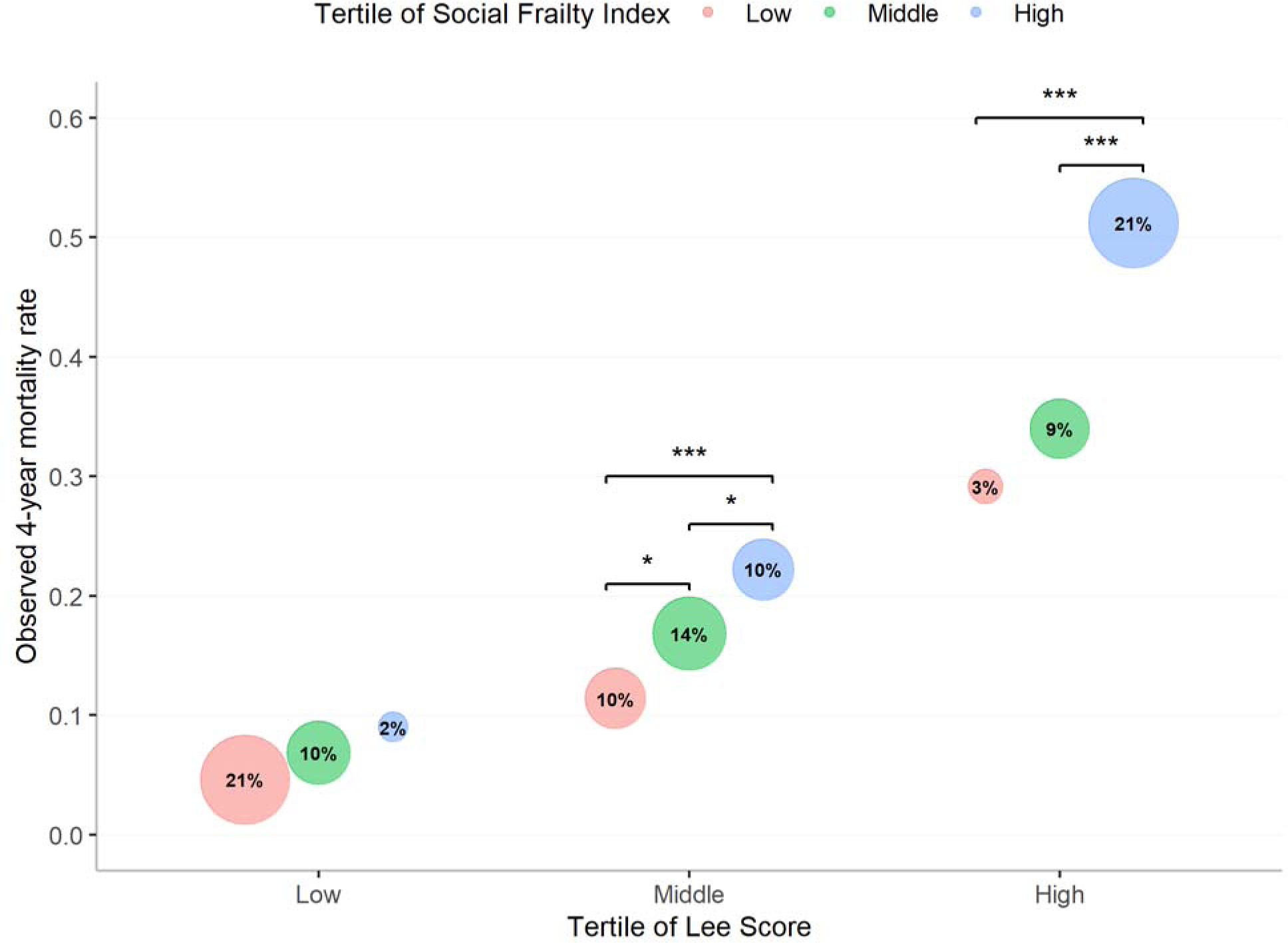
Observed mortality in 2012 Validation Cohort by Social Frailty and Lee Index The bubble chart compares observed mortality in the validation cohort by tertile of Social Frailty within tertiles of the Lee Index score, a comorbidity and function risk model. The area of each bubble is proportional to the total validation cohort that falls the specific group (e.g., 21% of the cohort has a low Lee Index score and low Social Frailty Index score). Significantly different values are highlighted by a bracket. *** p<0.001, ** p<0.01, * p<0.05. Results presented in tabular form in Appendix 7.

## DISCUSSION

Using a comprehensive social well-being survey of older adults, we developed and validated the Social Frailty Index, which predicts the risk of death over 4 years (**Figure 3**). We demonstrate that a small subset of social predictors can meaningfully stratify mortality risk in a nationally representative cohort of older adults. Further, the Social Frailty Index improves risk stratification beyond the Charlson score, a commonly used medical comorbidity model, and the Lee Index, a commonly used mortality prediction model. Where mortality prediction is important for older adults, this study demonstrates social risk factors represent an important and often unaccounted for risk domain.

**Figure 3:**
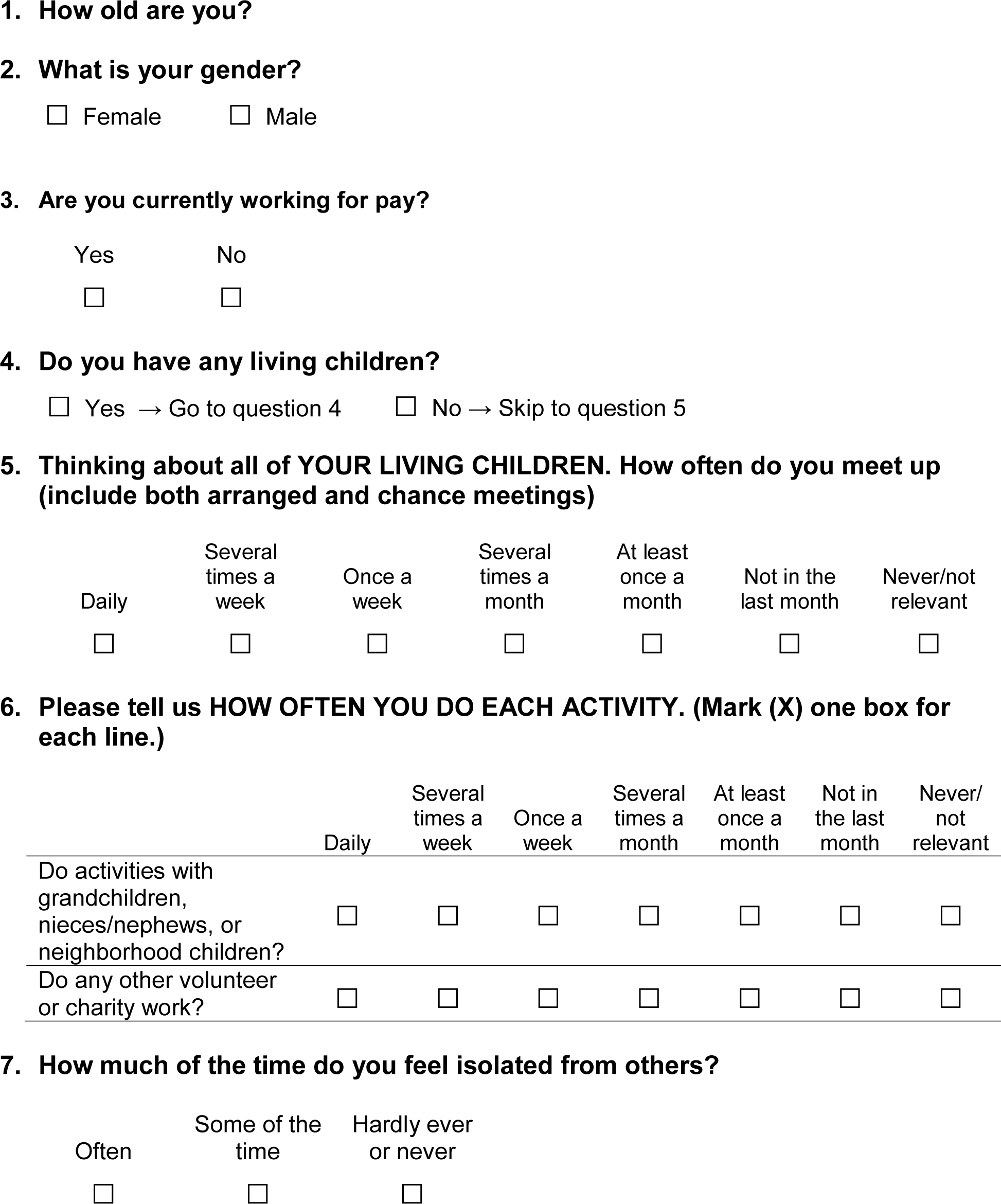
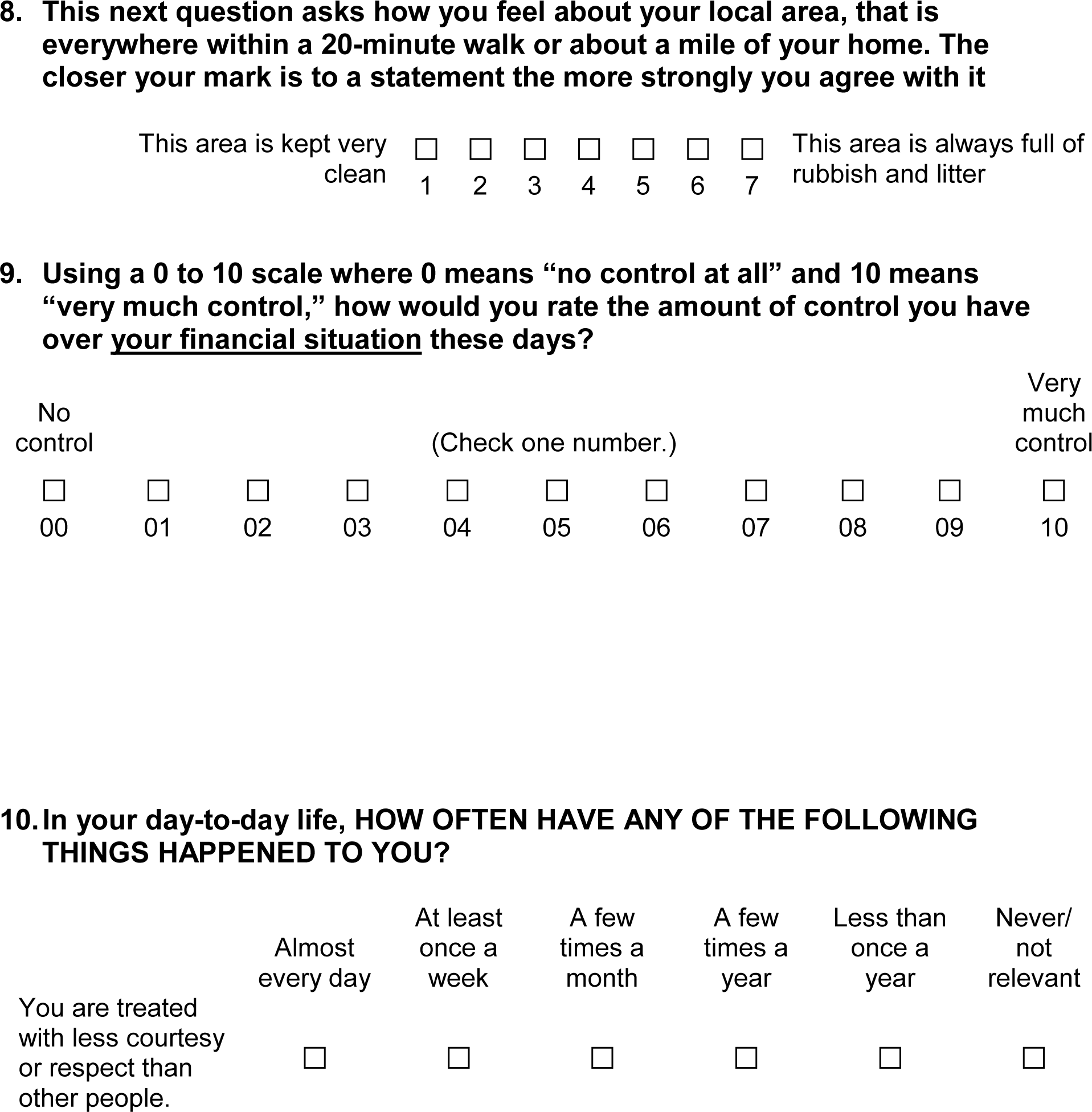
Social Frailty Index instrument. An online version is available at https://sachinjshah.shinyapps.io/Social_Frailty_Index/

This study builds on prior work incorporating social risk factors into prediction models in at least two noteworthy ways. First, we build on a wealth of studies establishing that social factors predict mortality by distilling the many hundreds of known social risk factors into an efficient summary index of social risk. The resulting Social Frailty index is a parsimonious model drawn from a comprehensive survey that can reasonably be exported. Second, we show that a comorbidity score alone is insufficient for risk stratification. We found social risk and medical comorbidities risk are not well correlated. Thus, when added to the Charlson model or Lee Index, the Social Frailty Index meaningfully recasts the predicted risk for a substantial number of individuals.

Where risk stratification is important, these results support supplementation of traditional prediction models with the Social Frailty Index.

There are several applications of the Social Frailty Index in medical research and clinical care. Collection in prospective observational studies is a natural use case for the index. Prognostic adjustment is central to observational research. The study of any risk factor hinges on accounting for the baseline risk differential in those with and without the prognostic factor in question. Current approaches rely heavily on medical comorbidities for prognostic adjustment. Adding the Social Frailty Index would help account for an acknowledged, but challenging to measure risk domain. In interventional studies, investigators may seek to understand if the effect of intervention differs by social frailty. Additionally, this instrument may be used in conjunction with existing models where mortality prediction is used to guide the clinical care of older adults. For instance, the Social Frailty Index may be used with legacy mortality prediction models when addressing advanced care planning or assessing the risk and benefits of screening procedures in older adults.

Additionally, the Social Frailty Index can address policymakers and health care delivery organizations’ need for accurate risk adjustment. For policymakers, quality measurement in health care turns on accurate baseline risk measurement. In a 2017 report, the National Academy of Medicine detailed the importance of including social risk factors in comorbidities-based risk models.^27^ However, social factors have yet to be included in quality measures, partly because it is not clear which measures to use.

Accountable Care Organizations commonly use claims and electronic health record data to identify patients for interventions like intensive care management. However, these methods do not adequately capture patients’ social dimensions, resulting in misidentification.^28, 29^ The Social Frailty Index could address significant gaps in quality measurement and population health.

The study design and data have limitations that are important to consider when interpreting the results. First, the risk factors in the Social Frailty Index are not necessarily causal; that is, it should not be taken to mean that addressing the risk factors identified in the model will reduce mortality risk. Additionally, the goal of this endeavor was to identify a small subset of social factors that best capture social risk. Thus, the absence of a putative factor in the index does not imply it was not predictive, rather that it was possible to capture the prognostic value of that risk factor across the study population through the risk factors already included in the model. Finally, in this study, the development and validation cohorts were separated by two years, a structure that lends credibility to the generalizability of the Social Frailty Index.^30^ Future validation outside of the Health and Retirement Study will prove helpful in characterizing the robustness of the Social Frailty Index. In particular, validation studies should focus on populations typically underrepresented in survey studies, like seniors living in poverty or racial and ethnic minorities such as Asian Americans.

In summary, the Social Frailty Index is a short survey that uses social risk factors to estimate the 4-year mortality risk in adults 65 years and older. The 10-item index obtained by patient report can be used to assess mortality risk and the risk of disability and prolonged nursing home stays. The model improves upon existing risk prediction tools and has clinical, population health, and research applications.

## Data Availability

Researchers can apply to the Health and Retirement Study (https://hrs.isr.umich.edu/) to access the data used in this study.

## ACKNOWLEDGMENTS

We would like to thank Dr. Leah Karliner and Dr. Michael Steinman for their participation in the advisory panel for this study.

## Author contributions

Dr. Shah had full access to all of the data in the study and takes responsibility for the integrity of the data and the accuracy of the data analysis. All authors listed have contributed sufficiently to the project to be included as authors, and all those who are qualified to be authors are listed in the author byline.

## Ethical review

The University of California, San Francisco, Committee on Human Research approved analysis for this study and waived the requirement for patient consent (Institutional Review Board No. 16-19185).

## Appendix

**Appendix 1:**
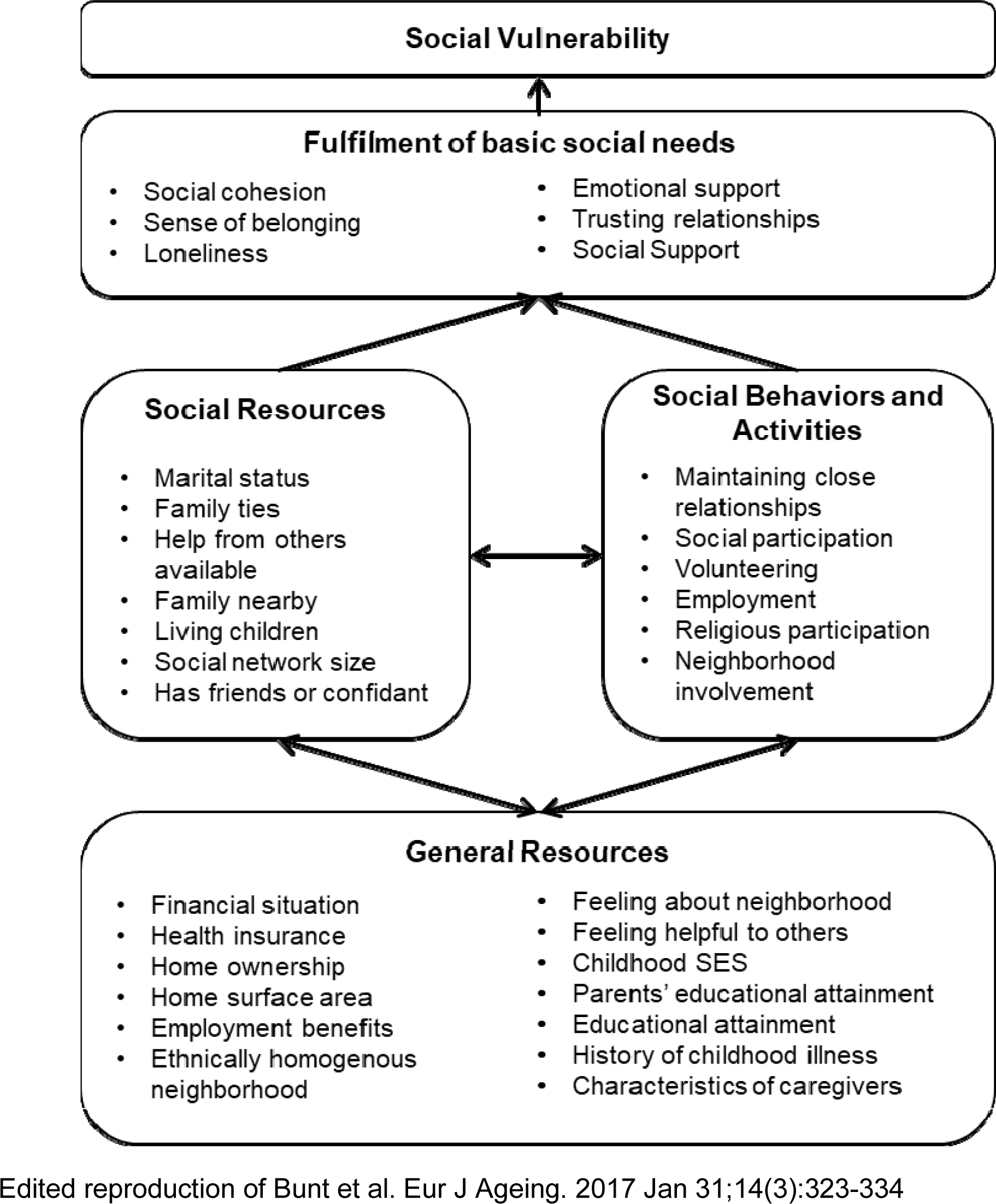
Social Frailty in Older Adults Framework

**Appendix 2:**
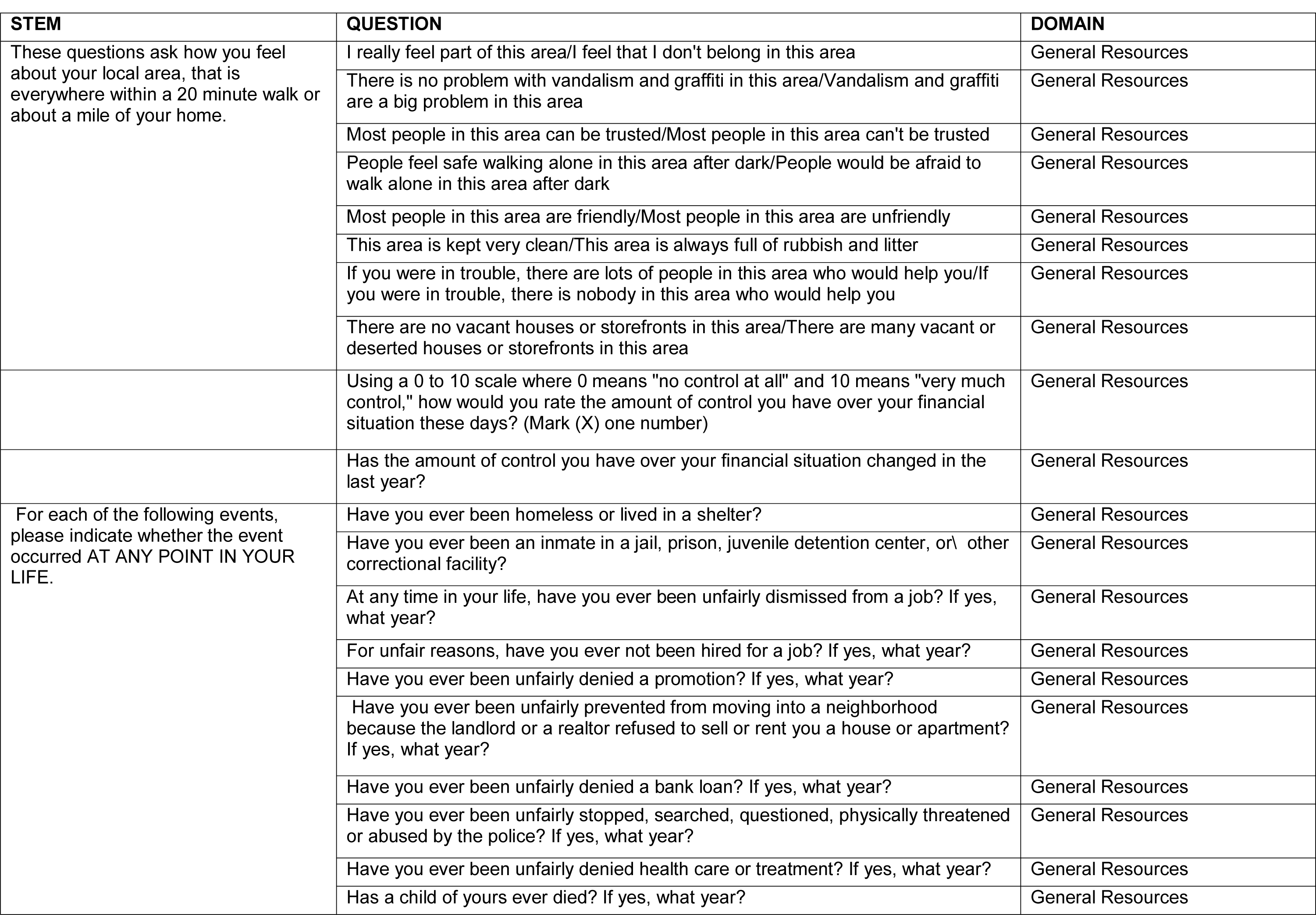
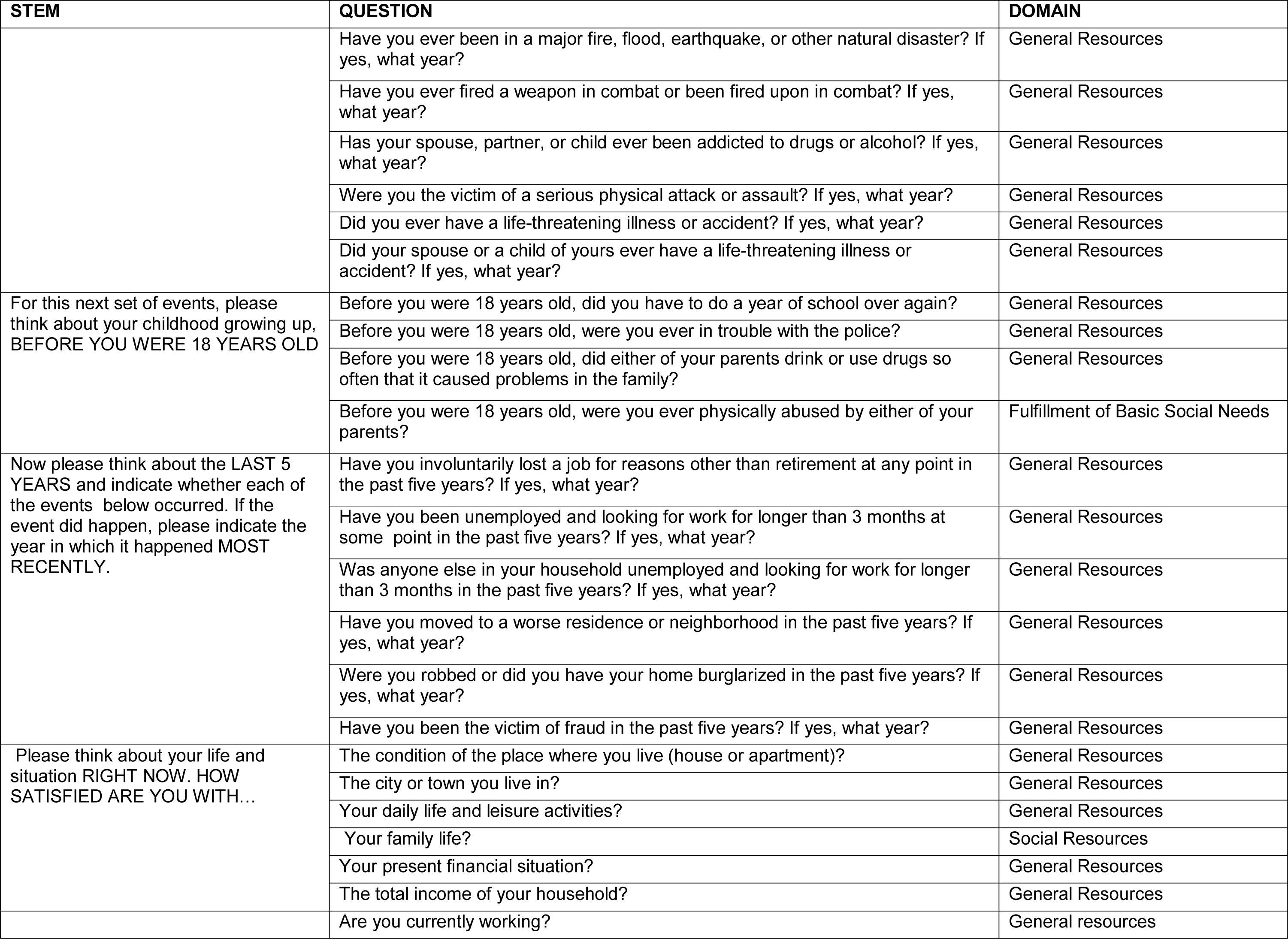
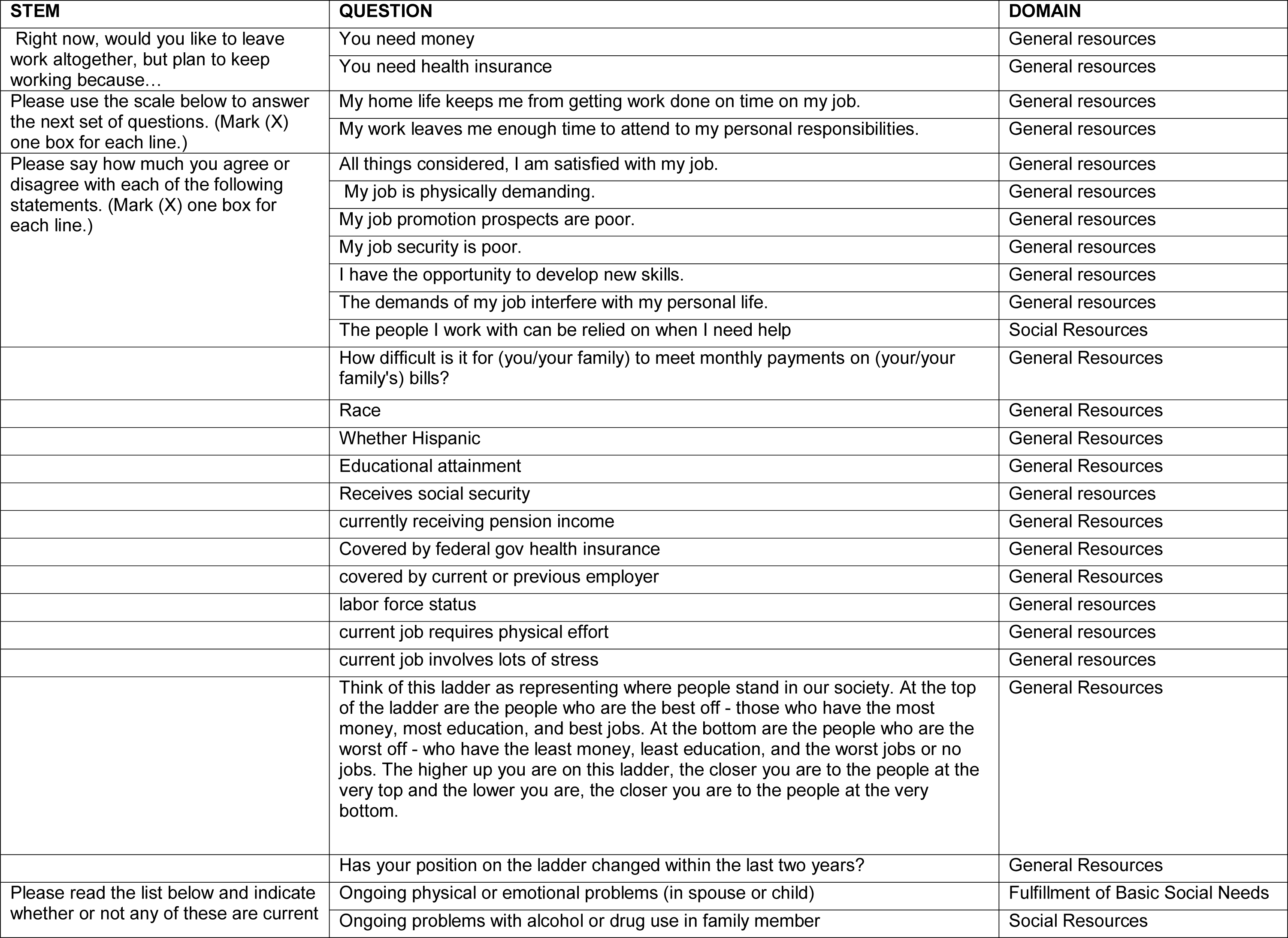
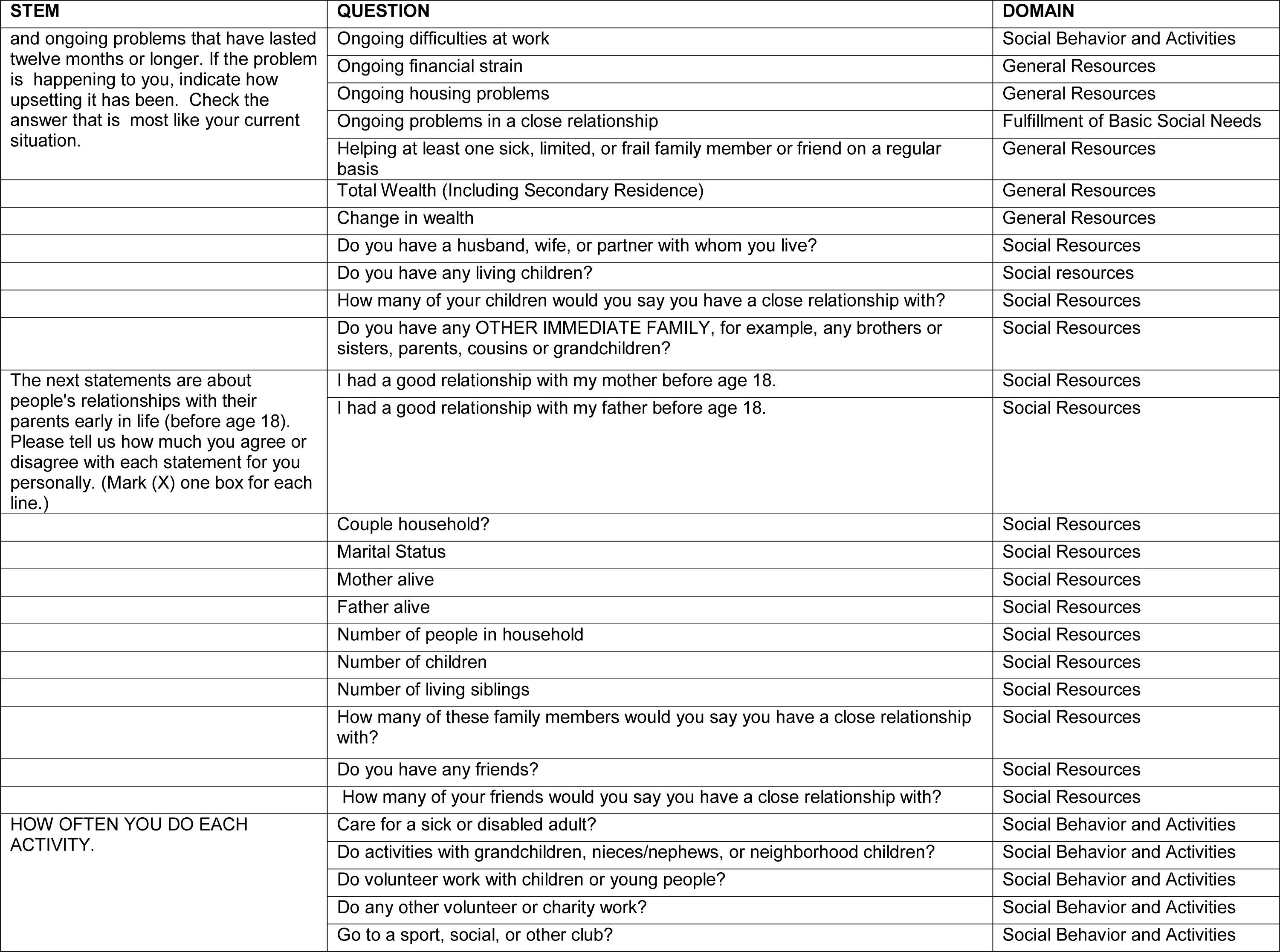
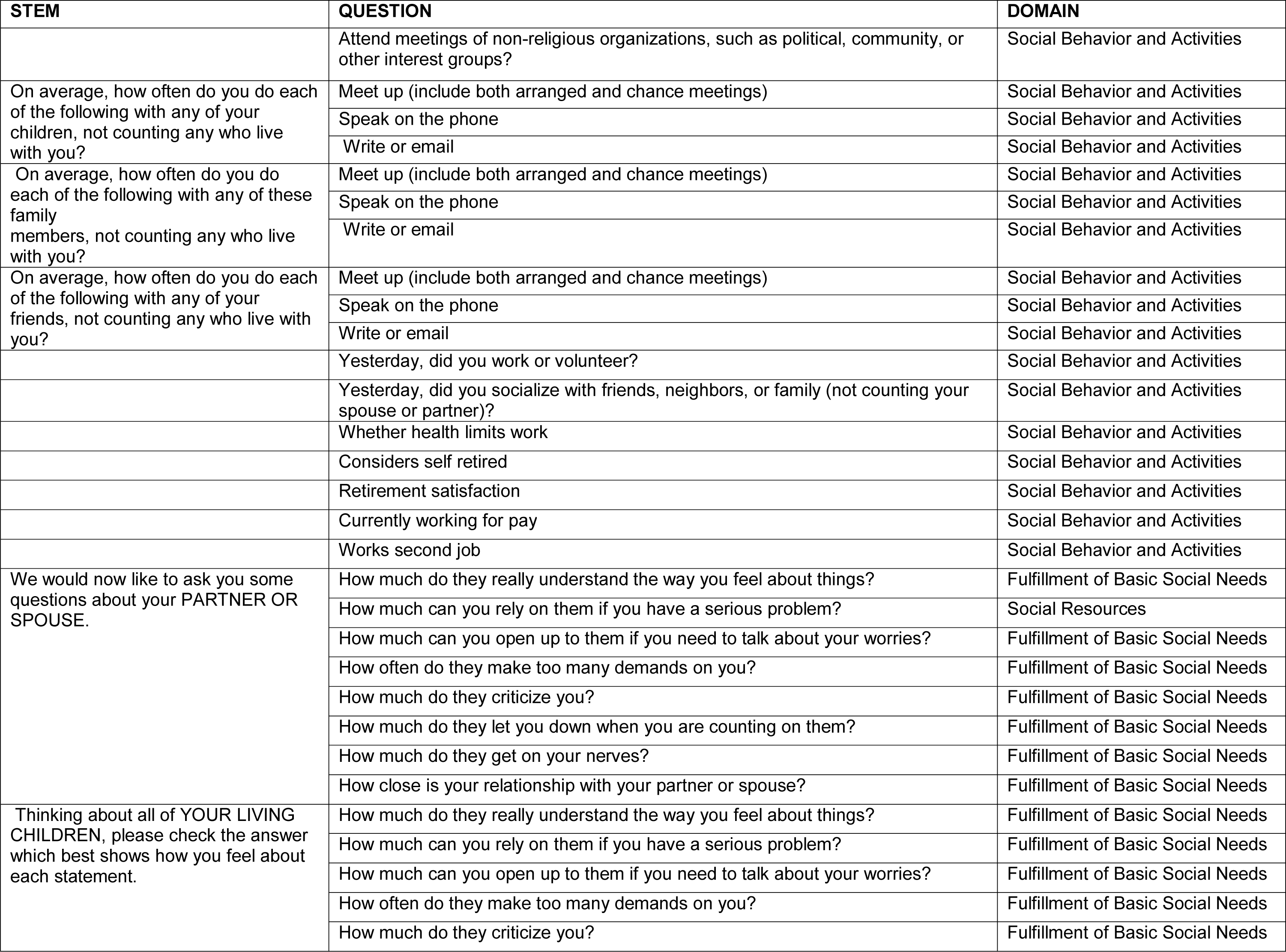
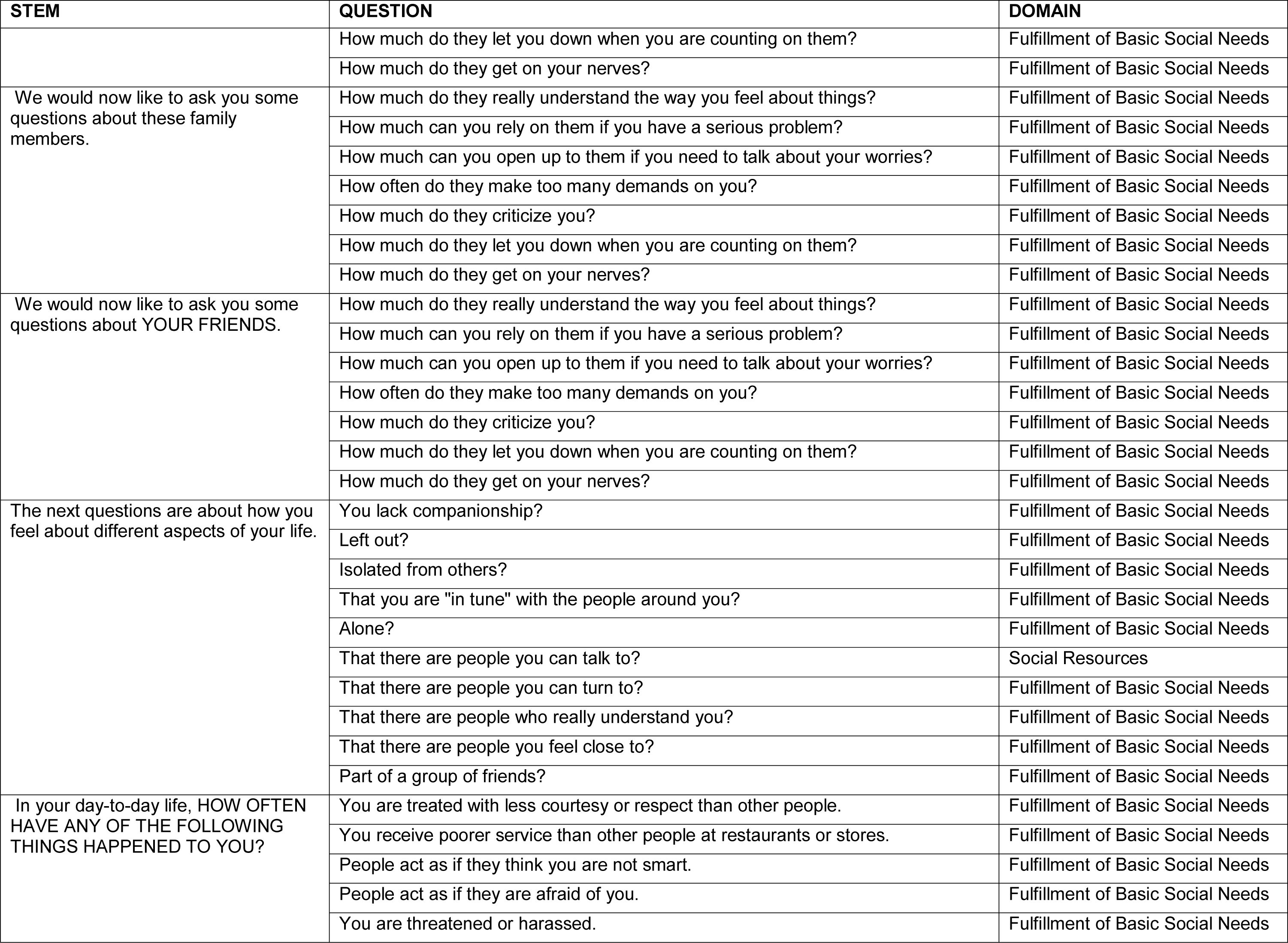
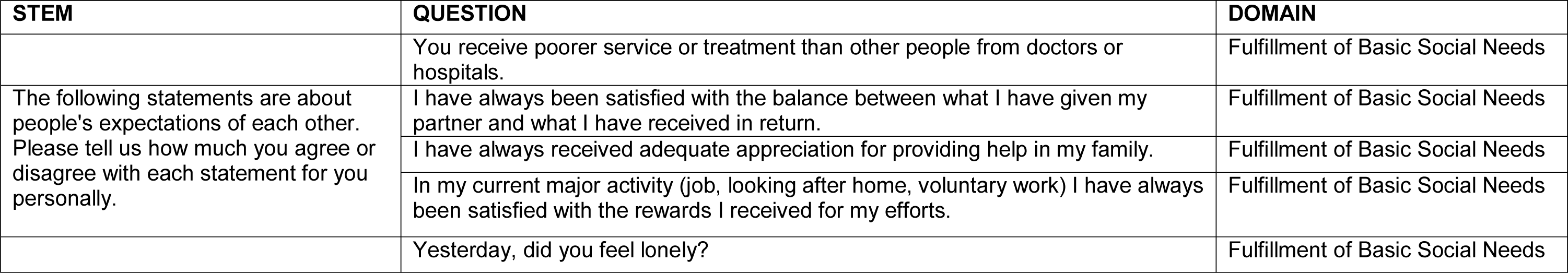
Candidate social predictors

**Appendix 3:**
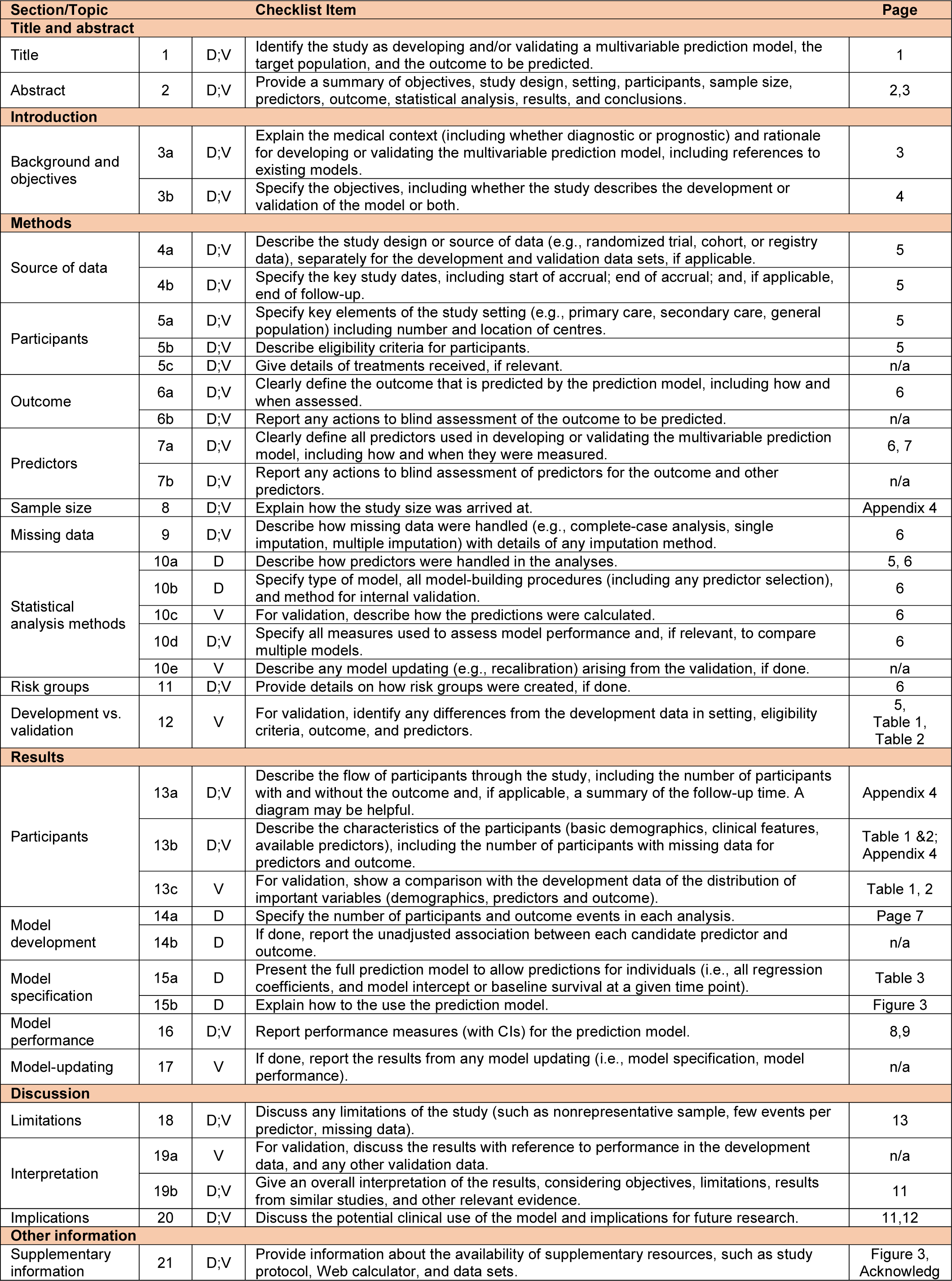
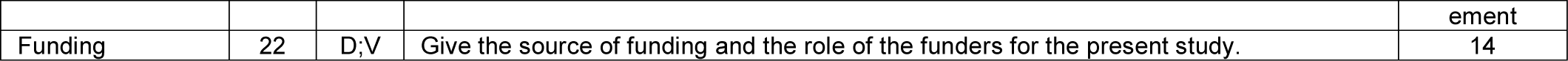
TRIPOD CHECKLIST

**Appendix 4:**
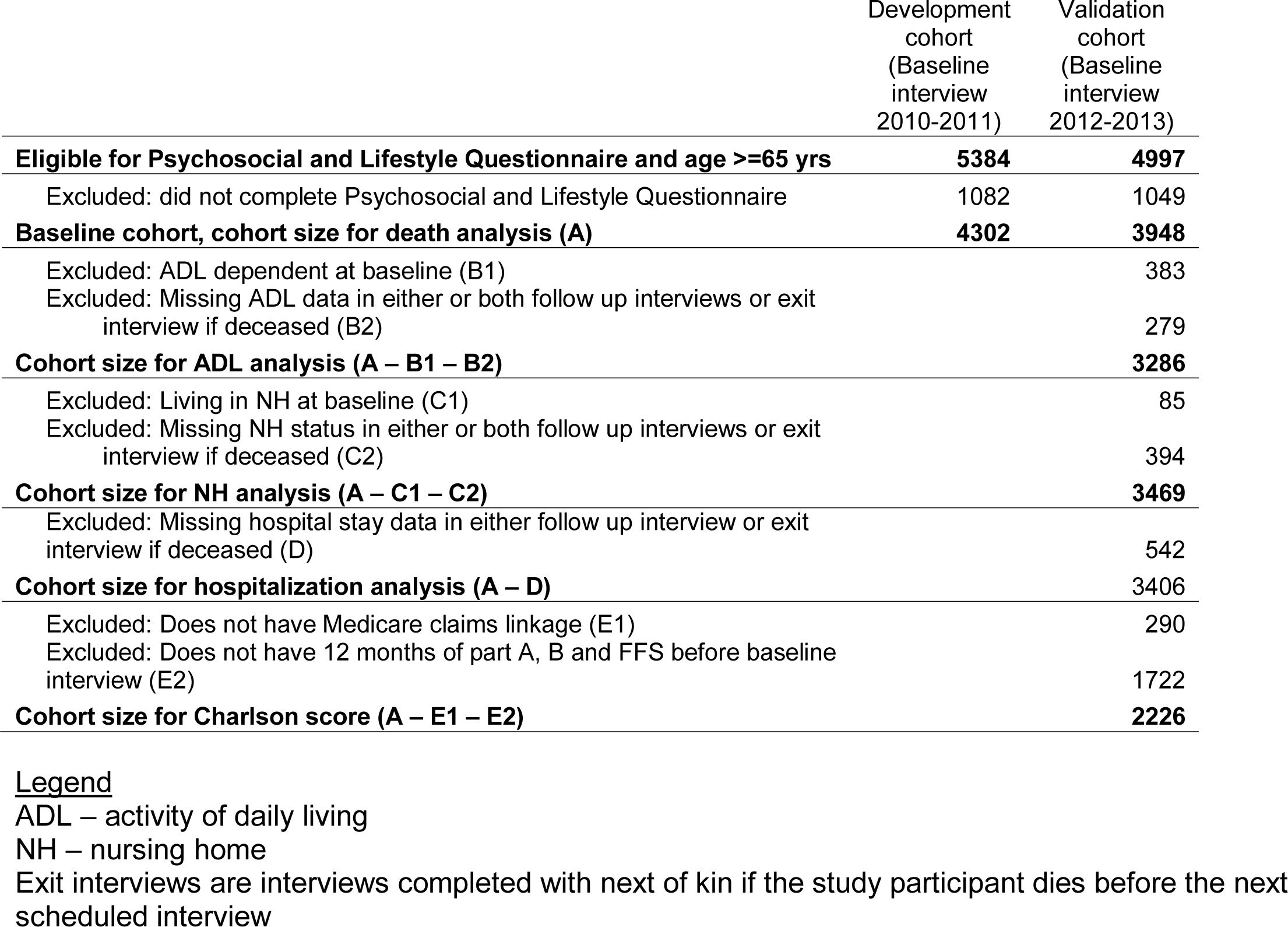
Cohort flow table

**Appendix 5:**
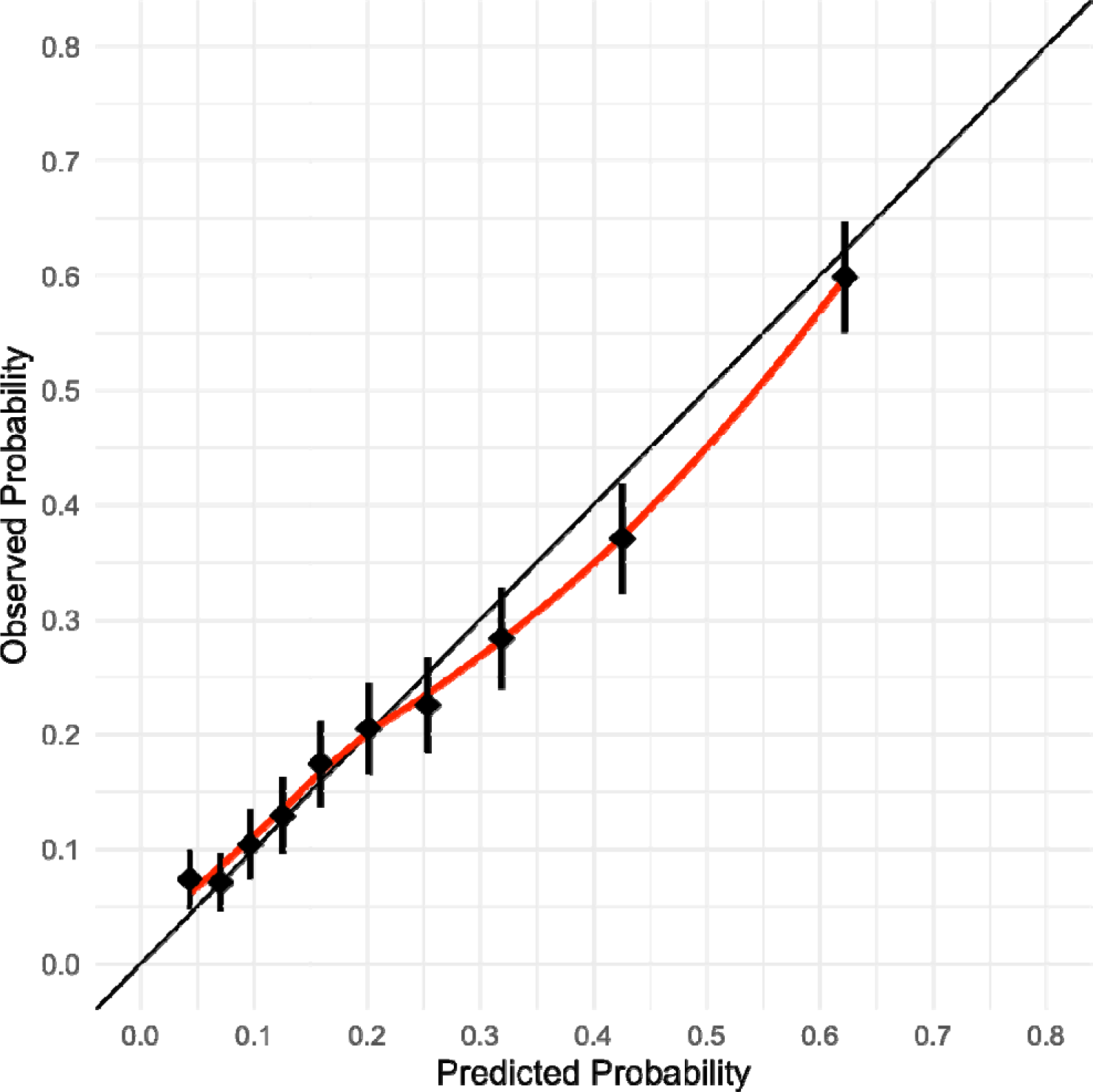
Calibration of Social Frailty Model in 2012 validation cohort

**Appendix 6:**
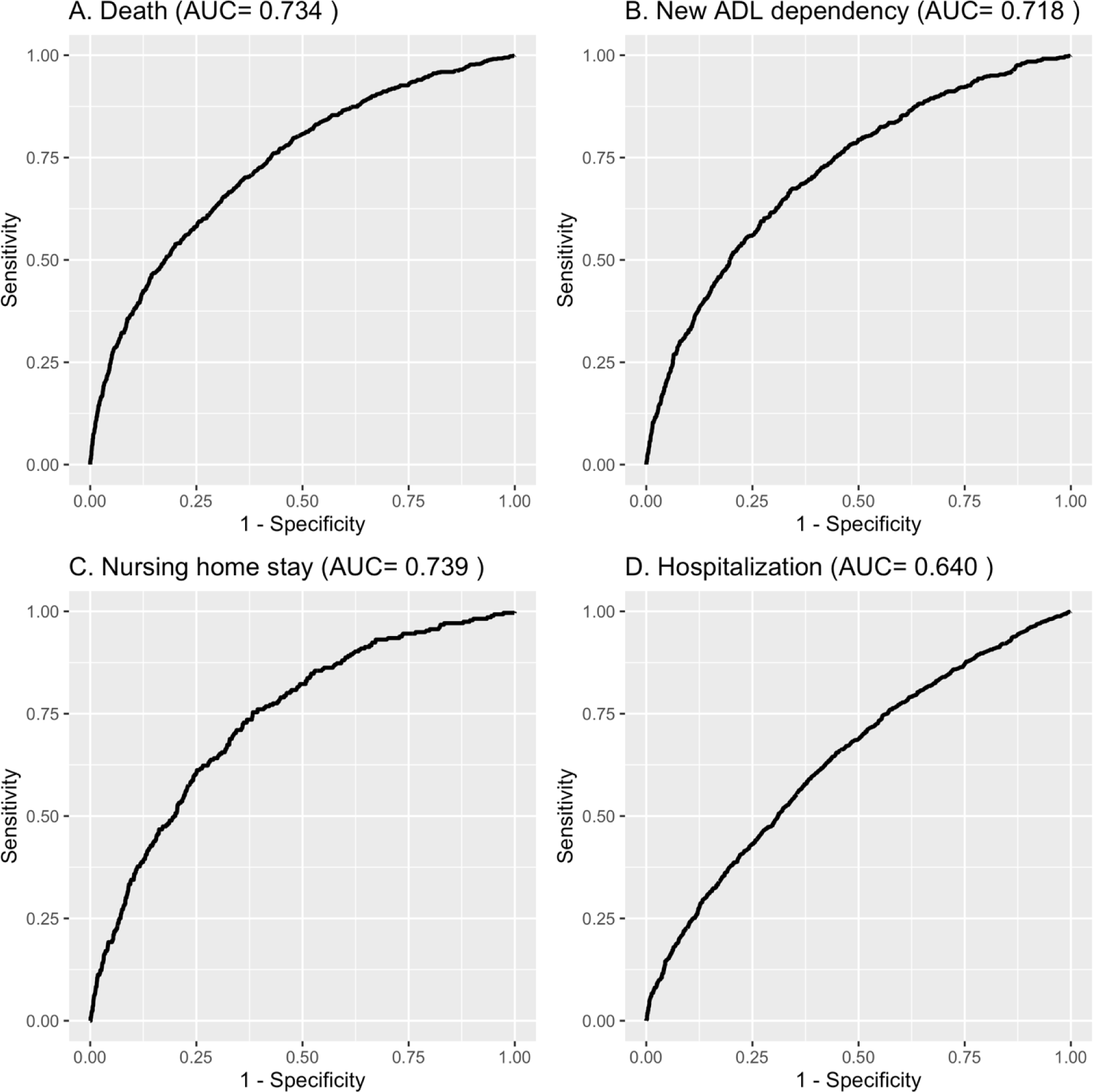
Figure Receiver Operator Curve for the Social Frailty Index to predict (A) Death, (B) New ADL dependence (C) Nursing home stay, and (D) Hospitalization in the 2012 Validation cohort

**Appendix 7:**
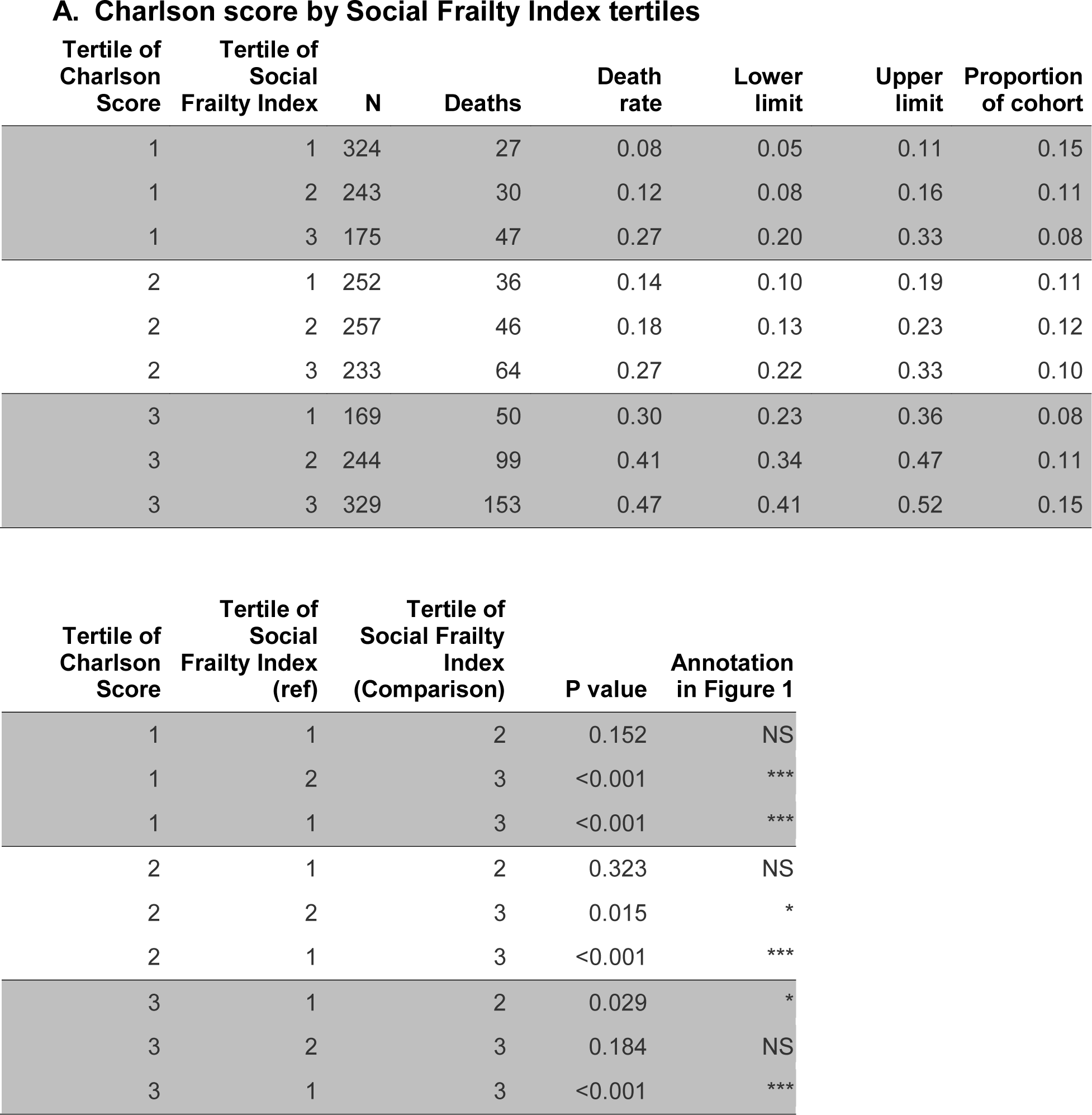
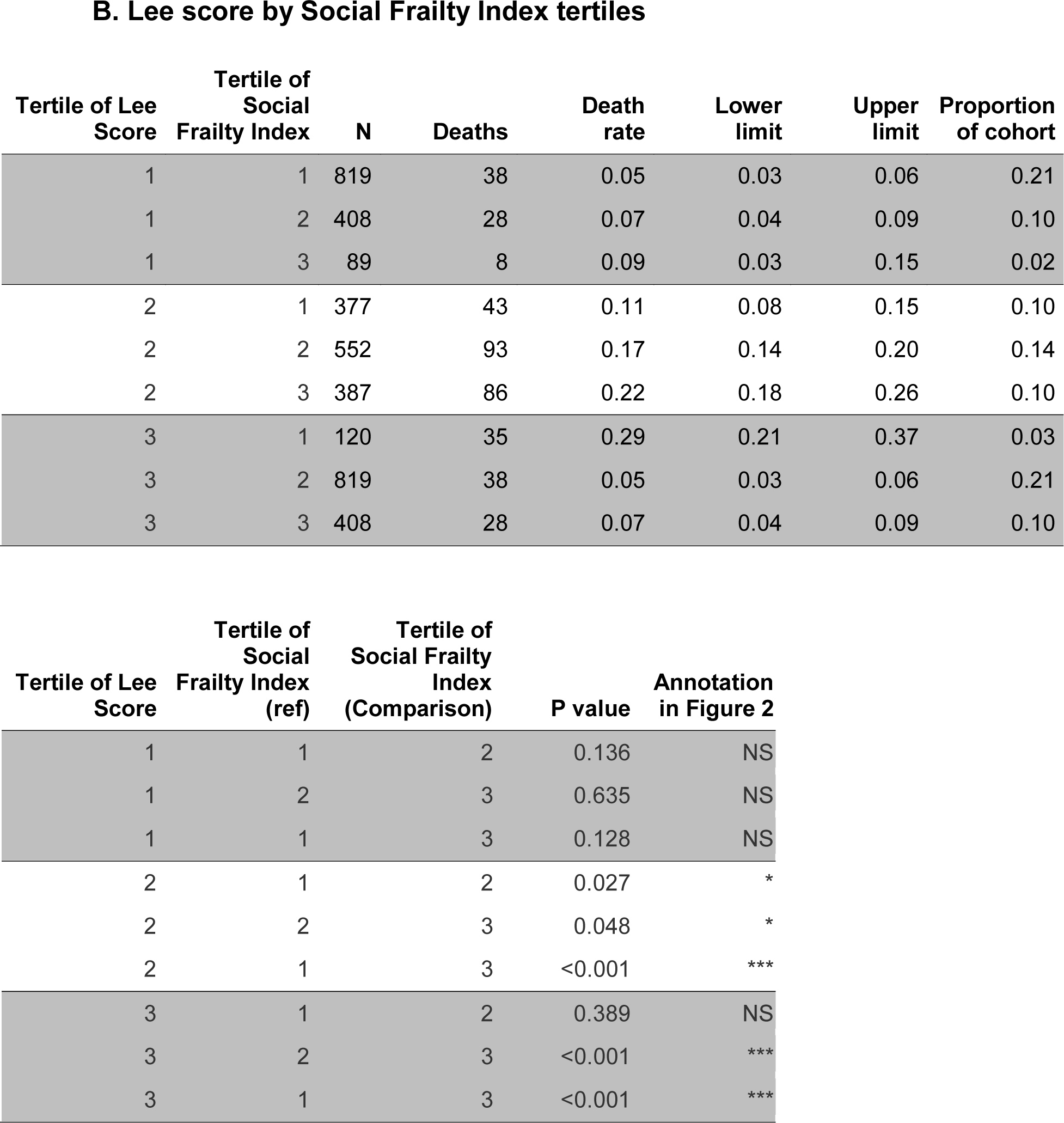
Tabular results of **Figure 1** and **Figure 2**

